# Cerebral small vessel disease burden in acute ischemic stroke and the role of physical activity: cross-sectional study

**DOI:** 10.1101/2025.04.23.25325707

**Authors:** Andreas Gammelgaard Damsbo, Rolf Ankerlund Blauenfeldt, Sigrid Breinholt Vestergaard, Niels Lech Pedersen, Kim Morgenstjerne Ørskov, Mette Foldager Hindsholm, Arzu Bilgin-Freiert, Claus Ziegler Simonsen, Søren Paaske Johnsen, Rikke Beese Dalby, Grethe Andersen, Janne Kaergaard Mortensen

## Abstract

**Background:** Cerebral small vessel disease (cSVD) is a major cause of stroke and cognitive decline. While classical cardiovascular risk factors are well-established contributors to overall cSVD burden, the effect of physical activity (PA) is not fully understood. This study aims to investigate the association between PA and cSVD burden in patients with acute ischemic stroke (AIS).

**Methods:** This is a post hoc analysis using pooled data from patients enrolled in two randomized stroke trials. cSVD burden was quantified as presence of microbleeds, lacunes, white matter hyperintensities and atrophy on acute admission magnetic resonance imaging (MRI) with a higher score corresponding to a higher overall cSVD burden (range 0-4). Pre-stroke PA was assessed by questionnaire on admission and grouped by quartiles (low quartile is low PA level). Ordinal logistic regression analyses were used to evaluate the association of PA and cSVD burden.

**Results:** A total of 762 patients with AIS were included. The median (IQR) age was 71 (62, 79) and 279 (37 %) were females. The proportion of patients with a cSVD score of 0 were 26 %, 38 %, 43 % and 57 %, respectively, through lowest to highest PA quartile. In the multivariable analysis the odds ratios for a higher cSVD score were lower in fourth PA quartile 0.63 (confidence intervals: 0.43 to 0.93), third 0.86 (0.57 to 1.29) and second 0.56 (0.36 to 0.87) quartile compared to the first quartile.

**Conclusion:** Among patients with AIS, a higher PA level was independently associated with a lower cSVD burden. This indicates a protective effect of PA on cSVD burden beyond modification of vascular risk factors.

## Introduction

Cerebral small vessel disease (cSVD) is one of the main causes of stroke but also the most important modifiable risk factor for, and second leading cause of, dementia.^1,2^

The cSVD is a multifactorial disease entity comprised of several underlying pathologic processes affecting the global cerebral circulation.^3^ Several features on magnetic resonance imaging (MRI) have been identified as biomarkers of cSVD, including cerebral microbleeds, chronic lacunes of presumed vascular origin, deep white matter hyperintensity (WMH) and atrophy, which all have been recommended as measures of the overall cSVD burden.^3,4^

Physical activity (PA) is appreciated as an important element in preventing and reducing cardiovascular risk factors such as hypertension and diabetes and reducing the negative effect of smoking,^5,6^ all risk factors shared by cSVD, stroke and dementia. Guidelines on stroke and dementia prevention include PA,^2,7^ but the association between cSVD and PA is still not clear.^8–10^

The assumed effect of PA on cSVD is multifaceted. PA may reduce classical cardiovascular risk factors mentioned previously, modify the effect of these cardiovascular risk factors, and may act directly on cerebral vessels by improving endothelial function, and lowering circulating fibrinogen levels.^11–14^

A higher level of PA has been associated with lower cSVD burden in the general population and among patients with cognitive impairment,^10,14–16^ while studies describing this association among patients with stroke are scarce.

We aimed to investigate how PA correlates to the burden of cSVD among patients with acute ischemic stroke (AIS) by evaluating the distribution of cSVD burden across different levels of pre-stroke PA, adjusted for common cardiovascular risk factors.

## Methods

### Study population

This study is a post-hoc, cross-sectional study on baseline MRI data from patients with AIS, pooled from two large randomized, clinical trials: the Efficacy of Citalopram Treatment in Acute Ischemic Stroke (TALOS) and the Remote Ischemic Conditioning for Acute Stroke (RESIST) trials.^17,18^

The TALOS trial included patients with first-time stroke within seven days after stroke onset who were randomized to citalopram or placebo treatment.^17^ In the RESIST trial, patients presenting with a prehospital putative acute stroke within four hours of symptom onset were randomized in the ambulance to ischemic conditioning treatment or sham treatment.^18^

Patients included in this current study were all admitted to the comprehensive stroke centre at Aarhus University Hospital, Aarhus, Denmark between 2013-2022. All received a final diagnosis of AIS and had a pre-stroke PA assessment available as well as an acute MRI scan. The acute MRI protocol conducted included diffusion weighted imaging (DWI) sequence, a fluid attenuated inversion recovery (FLAIR) sequence as well as T2*-weighted or susceptibility-weighted imaging (SWI) sequences.

### SVD annotation and scoring

Based on the available MRI sequences, cSVD biomarker annotation was evaluated for each subject by two independent, trained assessors according to the Standards for Reporting Vascular Changes on Neuroimaging (STRIVE) definitions.^19^ In case of disagreement, consensus scoring was performed by a third independent assessor. Assessors were blinded to clinical patient data and other assessors’ evaluations.

MRI biomarkers of cSVD have been combined in different ways for overall scoring systems of the cSVD burden in prognostication of long-term outcome^20,21^ and cSVD progression,^9^ but no standardized scoring systems exists to visually quantify the overall burden of cSVD.

In this study, the cSVD score was calculated by allocating one point for presence of each of the following: cerebral microbleeds(≥1), lacunes of presumed vascular origin (≥1), white matter hyperintensity (Fazekas score 2-3 in deep white matter) and global cerebral atrophy (Global Cortical Atrophy score 2-3).^22,23^ Thus, the cSVD score had a range of 0-4 with a higher score indicating more severe cSVD burden. Cerebral microbleeds were evaluated on T2* or SWI sequences, while the other cSVD markers were evaluated based on their presence on the FLAIR sequence (Figure 1).

**Figure 1.**
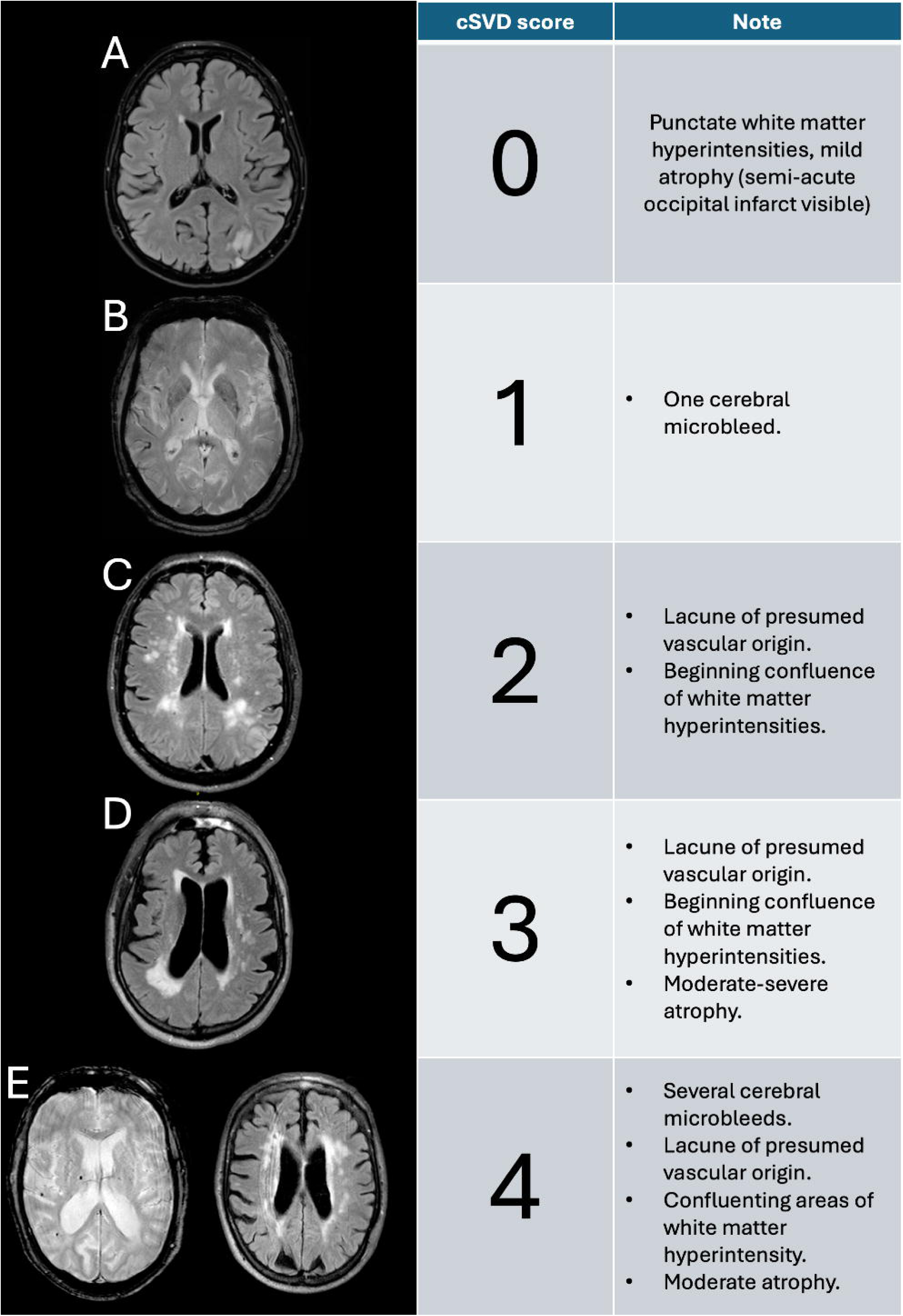
Example scores of the simplified cSVD score used from different subjects. Panel B and E1 are from T2* weighted sequences, the other panels are all FLAIR sequences.

All MRI scans were performed on scanners with a magnetic field strength of 1.5 or 3 Tesla.

### Clinical data

All clinical data was retrieved from the Danish Stroke Registry.^24^ The registry includes demographic and lifestyle data and we included data on age and sex as well as smoking habits (never, former, active), alcohol consumption (above 7 or 14 units of alcohol weekly for females and males, respectively), cohabitation status (living alone) and pre-stroke functional status (assessed by the modified Rankin Scale (mRS)), and data on hypertension, diabetes, prior cerebral ischemic event (AIS or transitory ischemic attack (TIA)), previous myocardial infarction (MI) and atrial fibrillation (AFib). Having diabetes, hypertension or AFib was defined by either receiving treatment, having a current diagnosis at the time of stroke admission, receiving a diagnosis or starting treatment during the hospital stay. All other characteristics were registered on stroke admission.

### Physical activity

Pre-stroke PA was defined by the PA level during the past 7 days prior to stroke. Pre-stroke PA was assessed using the Physical Activity Scale for the Elderly (PASE) questionnaire on trial enrolment.^25^ PASE is a validated 12-item questionnaire on overall PA including a variety of everyday activities including work, leisure time, household, and sports activities. The different items are summed through a weighted algorithm to give a score ranging from 0 to above 700, with a higher score indicating a higher level of PA. In a cohort of sedentary, older individuals (mean age 66.5) a median PASE score of 123 was found.^25^ The PASE questionnaire was completed by the patient or next of kin, and if necessary, with the assistance of study personnel. In the analyses, the PA score is divided into quartiles, with the first quartile corresponding to the lowest PA scores.

### Statistics

Baseline characteristics of included patients are presented in a table and the distribution of cSVD scores stratified by PA level is visualized in stacked bar plots. Comparisons of baseline factors were performed using the Kruskal-Wallis rank sum test and the Pearson’s Chi-squared test where relevant. All calculations were performed using 95 % confidence intervals (CI).

The correlation of PA level and cSVD burden was evaluated using ordinal logistic regression to evaluate the odds of a higher cSVD burden score with PA level as the main exposure.

In the regression analyses, age and sex were considered confounders of the association between PA and cSVD, while all other available covariables were considered potential mediators. This led to different adjustment strategies for the regression analyses: an unadjusted, univariable model; a minimal, confounder-adjusted model adjusted for age and sex; and a multivariable model with all available variables included.

Sensitivity analyses were performed as regression analyses stratified for sex as well as on data filtered to only include patients with pre-stroke mRS of 0 and without previous ischemic events (AIS or TIA). Cases with incomplete data were excluded from the regression analyses.

## Results

In total, 762 patients with available MRI and PA data were included: 397 from the RESIST cohort and 365 from the TALOS trial. Overall, RESIST patients were older, had a lower PASE score, fewer smoked regularly, more had hypertension and/or previous TIA. The TALOS cohort did not include patients with previous AIS as these were first time stroke patients.

All baseline characteristics are presented in Table 1. Overall, median age was 71 (IQR: 62-79), 279 (37 %) were female, and median PASE score was 108 (IQR: 60-160). Complete data were present on 733 subjects. Patients in the lower PA quartiles were older and more were females, more had hypertension, diabetes, AFib and higher pre-stroke mRS scores compared to the higher quartiles.

**Table 1.** Baseline characteristics and cSVD score characteristics stratified by physical activity quartile 1-4 (lowest-highest). cSVD: cerebral small vessel disease; GCA: Global Cortical Atrophy; MI: Myocardial infarction; mRS: modified Rankin Scale; WMH: White matter hyperintensity.

| Characteristic | Overall N = 762 | Q1 N = 191 | Q2 N = 191 | Q3 N = 189 | Q4 N = 191 |
| --- | --- | --- | --- | --- | --- |
| Age, Median (IQR) | 71 (62-79) | 77 (67-84) | 74 (65-80) | 70 (63-76) | 64 (55-73) |
| Female sex, n (%) | 279 (37%) | 85 (45%) | 89 (47%) | 58 (31%) | 47 (25%) |
| Living alone, n (%) | 203 (27%) | 70 (37%) | 54 (29%) | 45 (24%) | 34 (18%) |
| Missing, n | 4 | 1 | 2 | 1 | 0 |
| Smoking, n (%) |  |  |  |  |  |
| never | 269 (36%) | 57 (31%) | 75 (40%) | 69 (38%) | 68 (36%) |
| current | 211 (28%) | 56 (31%) | 45 (24%) | 53 (29%) | 57 (30%) |
| prior | 262 (35%) | 70 (38%) | 66 (35%) | 62 (34%) | 64 (34%) |
| Missing | 20 | 8 | 5 | 5 | 2 |
| High alcohol consumption, n (%) | 69 (9.2%) | 19 (10%) | 19 (10%) | 20 (11%) | 11 (5.8%) |
| Missing, n | 15 | 6 | 4 | 3 | 2 |
| Hypertension, n (%) | 418 (55%) | 123 (64%) | 114 (60%) | 96 (51%) | 85 (45%) |
| Diabetes, n (%) | 85 (11%) | 27 (14%) | 27 (14%) | 19 (10%) | 12 (6.3%) |
| Previous ischemic event, n (%) | 97 (13%) | 30 (16%) | 24 (13%) | 27 (14%) | 16 (8.4%) |
| Atrial fibrillation, n (%) | 116 (15%) | 34 (18%) | 29 (15%) | 29 (15%) | 24 (13%) |
| Previous MI, n (%) | 58 (7.6%) | 11 (5.8%) | 13 (6.8%) | 19 (10%) | 15 (7.9%) |
| Pre-stroke mRS, n (%) |  |  |  |  |  |
| 0 | 605 (79%) | 121 (63%) | 150 (79%) | 159 (84%) | 175 (92%) |
| 1 | 89 (12%) | 29 (15%) | 22 (12%) | 23 (12%) | 15 (7.9%) |
| 2 | 61 (8.0%) | 35 (18%) | 18 (9.4%) | 7 (3.7%) | 1 (0.5%) |
| 3 | 7 (0.9%) | 6 (3.1%) | 1 (0.5%) | 0 (0%) | 0 (0%) |
| **cSVD score characteristics** |  |  |  |  |  |
| cSVD score, Median (IQR) | 1 (0-2) | 1 (0-2) | 1 (0-2) | 1 (0-2) | 0 (0-1) |
| cSVD score features |  |  |  |  |  |
| Microbleeds ($\geq 1$), n (%) | 149 (20%) | 42 (22%) | 37 (19%) | 47 (25%) | 23 (12%) |
| Lacunes ($\geq 1$), n (%) | 241 (32%) | 71 (37%) | 60 (31%) | 61 (32%) | 49 (26%) |
| Beginning-large confluent areas of WMH (Fazekas 2-3), n (%) | 243 (32%) | 78 (41%) | 69 (36%) | 58 (31%) | 38 (20%) |
| Moderate-severe global atrophy (GCA 2-3), n (%) | 221 (29%) | 98 (51%) | 53 (28%) | 47 (25%) | 23 (12%) |

Stratified by PA level, patients with a cSVD score of 0 represented 26 %, 38 %, 43 % and 57 % respectively, from the lowest to the highest PA quartile (Figure 2).

**Figure 2.**
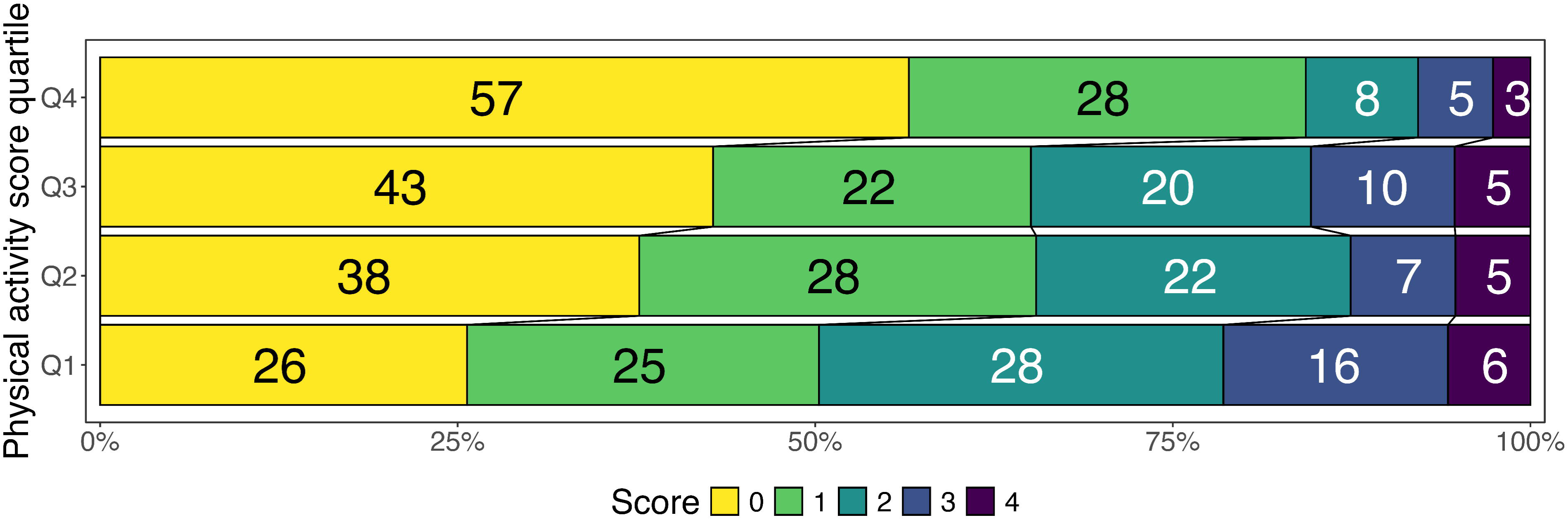
Relative distribution of cerebral small vessel disease (cSVD) burden score stratified by physical activity quartiles (Q1-4).

### Regression analyses

The ordinal regression models showed that a high PA level was associated with lower odds of a higher cSVD burden (Table 2).

**Table 2.** Ordinal regression models of cSVD burden score as main outcome with PASE score as the main exposure evaluating the odds ratio of higher cSVD score with a given PASE quartile compared to the first PASE quartile (lowest level of PA). CI: Confidence Interval; OR: Odds Ratio; PA: physical activity. A total of 733 subjects with complete data were included in the multivariable analysis. The minimally adjusted model is adjusted for age and sex. The multivariable model is adjusted for all available covariables.

|  | Univariable | Minimal | Multivariable |
| --- | --- | --- | --- |
| Characteristic | OR (95% CI) | OR (95% CI) | OR (95% CI) |
| Pre-stroke PA quartile |  |  |  |
| Q1 (lowest PA level) | — | — | — |
| Q2 | 0.57 (0.39 to 0.81) | 0.64 (0.44 to 0.93) | 0.63 (0.43 to 0.93) |
| Q3 | 0.52 (0.36 to 0.75) | 0.79 (0.53 to 1.16) | 0.86 (0.57 to 1.29) |
| Q4 (highest PA level) | 0.26 (0.17 to 0.37) | 0.51 (0.33 to 0.76) | 0.56 (0.36 to 0.87) |

The odds ratio (OR) for a higher cSVD score was 0.57 (CI: 0.39 to 0.81) for the second PA quartile, 0.52 (0.36 to 0.75) for the third and 0.26 (0.17 to 0.37) for the fourth (highest) PA quartile, compared to the first (lowest) PA quartile.

When adjusting for age and sex, a high PA level was still associated with lower odds of a higher cSVD burden. In the minimally adjusted model, the ORs (CIs) were 0.64 (0.44 to 0.93) for the second PA quartile, 0.79 (0.53 to 1.16) for the third, and 0.51 (0.33 to 0.76) for the fourth PA quartile, as compared to the first quartile.

When adjusting for all available covariables, the ORs (CI) were 0.63 (0.43 to 0.93) for the second PA quartile, 0.86 (0.57 to 1.29) for the third, and 0.56 (0.36 to 0.87) for the fourth PA quartile compared with the first quartile (Table 2). Results from univariate analyses of all considered covariables and all coefficients from the multivariable analyses are reported in the supplementary material (Supplementary table 1 and Supplementary figure 1).

Sensitivity analyses found the same overall pattern of correlation between cSVD score and PASE quartiles among males, females and in patients without pre-stroke functional disability (mRS score 0) or prior ischemic event (AIS or TIA) (Supplementary Table 2 and 3).

## Discussion

This was a cross-sectional study of the association between pre-stroke PA level and cSVD burden on acute MRI in a large cohort of patients with AIS.

Higher PA levels were associated with a lower cSVD burden, with patients in the highest PA quartile having 74% lower odds of an increased cSVD score. The minimally adjusted regression model adjusting for age and sex changed the point estimates of model coefficients, but the associations remained statistically significant. These results remained consistent, although reduced after further adjustments, indicating that the effect of PA on cSVD is mediated through other pathways than just modifying classical cardiovascular risk factors.

While an association between PA and cSVD has been shown previously,^9,15,16,26^ it has not been studied in a stroke population. One study included 590 older community-dwelling individuals.^15^ Similar to our results, they found an association between lower levels of PA and higher white matter hyperintensity volume as well as a higher number of microbleeds and lacunes. Another study on PA and progression of overall cSVD burden among 503 patients diagnosed with cSVD in a prospective design with a 9-year follow-up showed no association between baseline PA and neither overall cSVD progression nor progression on individual cSVD markers.^9^ PA assessment was based on a questionnaire estimating the average time spent on different activities per week during the last year, which may have increased the risk of recall bias, and diluted the results. The authors only used baseline PA in their analyses and did not take change in the PA level into account. The distribution of cSVD burden scores by PA level was also not reported, which limits the comparability to the current study.

Finally, a large study among 680 stroke-free Japanese males investigated the association of baseline daily step-counts and cSVD biomarkers after ∼7 years follow-up.^16^ A moderate level of PA was correlated to the lowest prevalence of cSVD, suggesting that a very high level of PA is not necessarily protective to the cerebral microcirculation or should be differentiated when addressing neuroprotectivity.

Limitations of this current study include, that the diagnoses of comorbid diseases were registered without any data on treatment status or severity, and did not include biomedical measures such as Haemoglobin A1C, lipid profile, or body mass index.

Furthermore, due to the nature of the study design, no clear conclusions on causality can be drawn without risking overlooking reverse causality, as patients suffering from cSVD may have gradually become more inactive before events leading to inclusion in a stroke cohort study. By including patients with stroke only, the cohort represents a group of patients at high cardio-vascular risk and not the general population. The relationship between PA and cSVD risk factors is complex, probably bidirectional, and clear inference of results from an observational or cross-sectional study like the present will never be conclusive.

We used the PASE questionnaire to assess pre-stroke PA upon stroke admission, which has been validated to reflect retrospective PA.^25^ To reduce the risk of recall bias, assessment was performed at a median of 2 days after stroke admission, and if necessary, study personnel assisted in the completion of the questionnaire. This study was limited to patients with PASE available, which may have resulted in a selection bias as patients with more severe strokes and more severe cSVD were not included. If higher PA decreases the risk of severe stroke and cSVD burden, this selection may have led to underestimating the association between PA and cSVD.

Only 37% of patients included were female, which reduces the generalizability of our findings. However, we did repeat our analyses stratified by sex and found no major difference in our results.

In this study we found a possible dose-response-like association with a higher PA level being associated with a lower cSVD burden. The relation between PA and overall health benefits is not perfectly clear. Some have argued for a proportional, dose-response-like association,^27^ while other studies have found a U-shaped relation between PA and cardiovascular risk with a plateau where a moderate PA level was associated with the greatest health benefits.^16,28^ Levels of PA in this current study may have been too low to show this possible U-shaped relation between PA and cSVD.

This large study on 762 patients with AIS showed a cSVD distribution indicating a proportional relationship of higher PA levels and lower cSVD burden. The regression analyses showed an independent association between a higher PA level and a lower cSVD burden. Though adjusting the model reduced the strength of the association, the results indicate that PA impacts cSVD through other pathways than just modification of cardiovascular risk factors. The benefit of a moderate-high PA lifestyle thus seems important, as already known for people in general, but also for patients with stroke to prevent or postpone cSVD development.

## Abbreviations

AIS: acute ischemic stroke
cSVD: cerebral small vessel disease
mRS: modified Ranking Scale
PA: physical activity
WMH: white matter hyperintensity
TALOS: Efficacy of Citalopram Treatment in Acute Ischemic Stroke
RESIST: Remote Ischemic Conditioning for Acute Stroke
DWI: diffusion weighted imaging
SWI: susceptibility-weighted imaging
FLAIR: fluid attenuated inversion recovery
MRI: Magnetic Resonance Imaging
TIA: transitory ischemic attack
MI: myocardial infarction
AFib: atrial fibrillation
PASE: Physical Activity Scale for the Elderly

## Acknowledgments

Support provided by the Steno Diabetes Center Aarhus (SDCA) which is partially funded by an unrestricted donation from the Novo Nordisk Foundation.

## Author contributions

AGD, RAB, RBD, GA, SPJ, and JKM contributed equally to conceptualization. AGD, RAB, SBV, NLP, KMØ, MFH, AB-F, JKM contributed equally to MRI assessments and CZS and GA made consensus assessments. AGD led formal analysis, and the original draft. GA led investigation, while SPJ, RAB, RBD and JKM led methodology. RBD and JKM supported original draft preparation. All authors contributed equally to manuscript revision.

## Statements and Declarations

### Ethical considerations

The TALOS trial was approved by the local institutional review board and the Committees on Health Research Ethics (ID: 1-10-72183-13). The RESIST trial was approved by Danish regional research ethics committees (ID: 1-10-72-97-17). The current study is approved by the Danish Data Protection Agency and the local Ethics Committee. Both studies were conducted in accordance with the principles of the Declaration of Helsinki.

### Consent to participate

All patients were included by written informed consent, or if necessary, consent from next of kin.

### Data availability

This study is based on sensitive data subject to EU legal restrictions. A subset of the data can however be made available from the corresponding author on a reasonable request.

