## Supplemental material for "Cerebral small vessel disease burden in acute ischemic stroke and the role of physical activity: cross-sectional study"

|  | Univariable | | | Multivariable | |
| --- | --- | --- | --- | --- | --- |
| **Characteristic** | **N** | **OR***^1^* | **95% CI***^1^* | **OR***^1^* | **95% CI***^1^* |
| Pre-stroke PA quartile | 762 |  |  |  |  |
| Q1 |  | — | — | — | — |
| Q2 |  | 0.57 | 0.39, 0.81 | 0.63 | 0.43, 0.93 |
| Q3 |  | 0.52 | 0.36, 0.75 | 0.86 | 0.57, 1.29 |
| Q4 |  | 0.26 | 0.17, 0.37 | 0.56 | 0.36, 0.87 |
| Age | 762 | 1.09 | 1.08, 1.11 | 1.09 | 1.07, 1.11 |
| Female sex | 762 | 1.19 | 0.91, 1.55 | 0.86 | 0.62, 1.18 |
| Living alone | 758 | 1.56 | 1.17, 2.07 | 1.05 | 0.75, 1.47 |
| Smoking | 742 |  |  |  |  |
| never |  | — | — | — | — |
| current |  | 1.01 | 0.73, 1.41 | 1.51 | 1.04, 2.20 |
| prior |  | 1.69 | 1.24, 2.32 | 1.36 | 0.97, 1.90 |
| High alcohol consumption | 747 | 1.45 | 0.94, 2.24 | 1.32 | 0.83, 2.07 |
| Hypertension | 762 | 2.90 | 2.22, 3.81 | 1.81 | 1.35, 2.45 |
| Diabetes | 762 | 1.50 | 1.01, 2.23 | 1.33 | 0.85, 2.07 |
| Previous ischemic event | 762 | 2.53 | 1.71, 3.75 | 2.06 | 1.36, 3.14 |
| Atrial fibrillation | 762 | 1.05 | 0.73, 1.50 | 0.56 | 0.37, 0.83 |
| Previous MI | 762 | 0.87 | 0.53, 1.42 | 0.52 | 0.30, 0.88 |
| Pre-stroke mRS | 762 |  |  |  |  |
| 0 |  | — | — | — | — |
| 1 |  | 1.58 | 1.06, 2.35 | 1.04 | 0.68, 1.59 |
| 2 |  | 3.06 | 1.90, 4.95 | 0.98 | 0.57, 1.68 |
| 3 |  | 2.97 | 0.92, 9.73 | 0.56 | 0.14, 2.28 |
| *^1^*OR = Odds Ratio, CI = Confidence Interval | | | | | |

Supplementary table 1 Univariable and multivariable ordinal regression analyses of higher small vessel disease burden. Includes univariable analyses of all considered covariables and includes all coefficients from the multivariable analyses. The multivariable analysis is the same as the main analysis as seen in Table 2 in the main article.

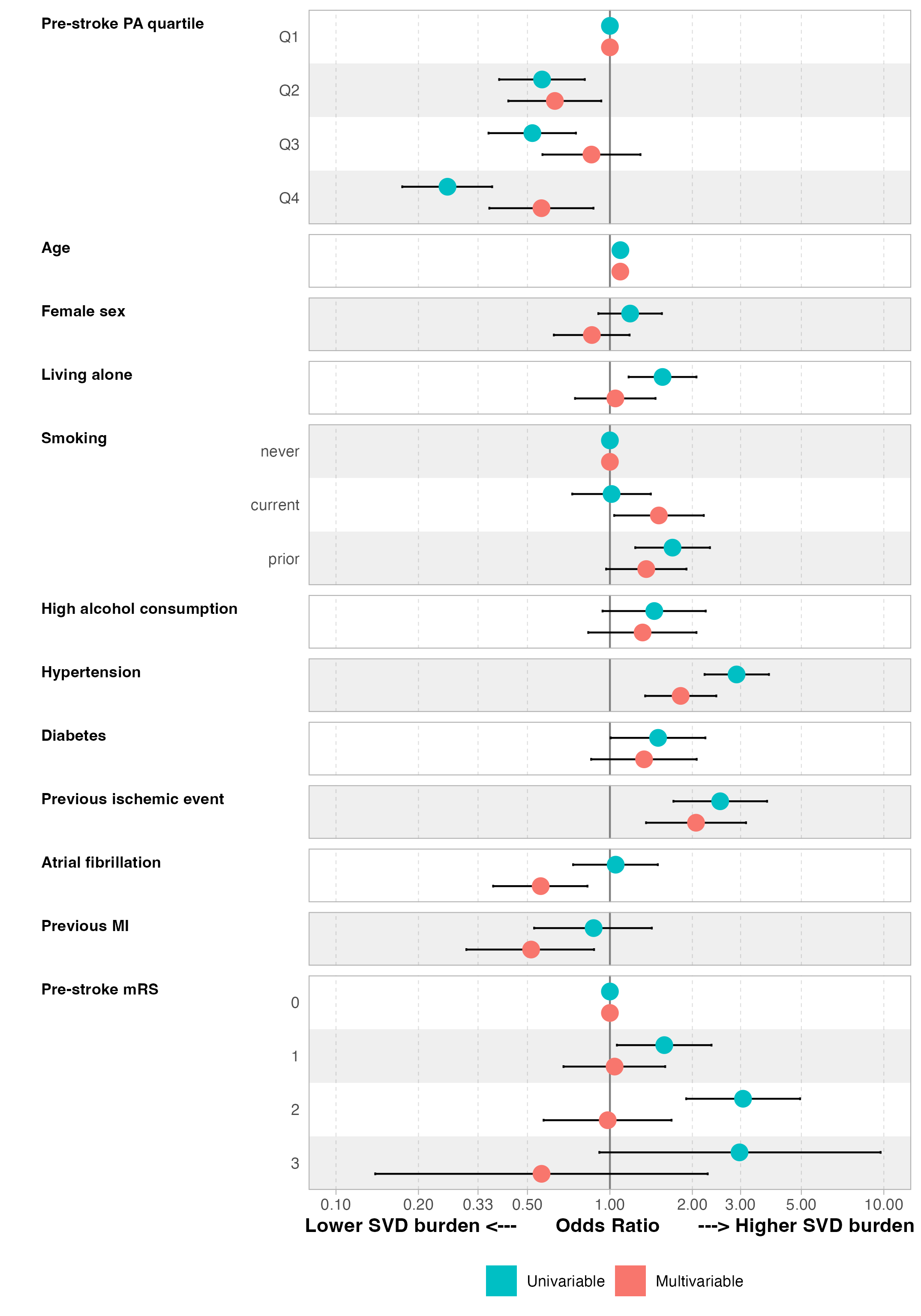

Supplementary figure 1 Univariable and multivariable ordinal regression analyses coefficients of higher small vessel disease burden plotted. Based on the values reported in Supplementary table 1.

|  | Univariable | Minimal | | Multivariable | |
| --- | --- | --- | --- | --- | --- |
| **Characteristic** | **OR** **(95% CI)***^1^* | **OR** **(95% CI)***^1^* | **p-value** | **OR** **(95% CI)***^1^* | **p-value** |
| **MALE** | | | | | |
| Pre-stroke PA quartile |  |  |  |  |  |
| Q1 | — | — |  | — |  |
| Q2 | 0.72 (0.44 to 1.17) | 0.73 (0.44 to 1.20) | 0.21 | 0.71 (0.42 to 1.20) | 0.20 |
| Q3 | 0.60 (0.38 to 0.96) | 0.73 (0.45 to 1.19) | 0.21 | 0.86 (0.51 to 1.44) | 0.57 |
| Q4 | 0.31 (0.19 to 0.49) | 0.49 (0.30 to 0.80) | 0.005 | 0.58 (0.34 to 0.97) | 0.040 |
| **FEMALES** | | | | | |
| Pre-stroke PA quartile |  |  |  |  |  |
| Q1 | — | — |  | — |  |
| Q2 | 0.42 (0.24 to 0.72) | 0.56 (0.32 to 0.98) | 0.044 | 0.51 (0.27 to 0.95) | 0.034 |
| Q3 | 0.43 (0.23 to 0.78) | 0.94 (0.48 to 1.84) | 0.86 | 1.02 (0.48 to 2.13) | 0.96 |
| Q4 | 0.19 (0.09 to 0.38) | 0.59 (0.27 to 1.29) | 0.18 | 0.61 (0.26 to 1.43) | 0.26 |

Supplementary table 2 Ordinal regression models of SVD burden score as main outcome stratified by sex with PASE score as the main exposure

|  | Univariable | Minimal | Multivariable |
| --- | --- | --- | --- |
| **Characteristic** | **OR** **(95% CI)***^1^* | **OR** **(95% CI)***^1^* | **OR** **(95% CI)***^1^* |
| Pre-stroke PA quartile |  |  |  |
| Q1 | — | — | — |
| Q2 | 0.59 (0.37 to 0.93) | 0.62 (0.39 to 1.00) | 0.65 (0.39 to 1.06) |
| Q3 | 0.55 (0.35 to 0.86) | 0.87 (0.54 to 1.41) | 1.03 (0.62 to 1.70) |
| Q4 | 0.30 (0.19 to 0.47) | 0.62 (0.38 to 1.02) | 0.78 (0.46 to 1.32) |
| *^1^*OR = Odds Ratio, CI = Confidence Interval | | | |
| Ordinal regression models of SVD burden score as main outcome with PASE score as the main exposure. | | | |

Supplementary table 3 Ordinal regression models of SVD burden score as main outcome with PASE score as the main exposure only including patients with pre-stroke modified Rankin Scale score of 0 (no disability) and no previous ischemic events (acute ischemic stroke or transient ischemic attack).
